# On the protective role of the blood vessels in glaucomatous damage: a transversal study

**DOI:** 10.1101/2021.03.17.21253785

**Authors:** Yaiza Cánovas-Serrano, Lorenzo Vallés-San-Leandro, Miguel Ángel Rodríguez-Izquierdo, Jerónimo Lajara-Blesa, Rafael López-Serrano

**Affiliations:** Health Sciences PhD program, Catholic University of Murcia (UCAM), Campus de los Jerónimos n°135, Guadalupe 30107, Murcia, Spain; Clinical Research Department, Vista Ircovisión, Murcia, Spain; Faculty of Health Sciences, Catholic University of Murcia (UCAM), Spain; Faculty of Economics, University of Murcia (UMU), Spain

**Keywords:** astrocytes, visual field, primary open-angle glaucoma, aqueous humor, blood vessel

## Abstract

**Purpose:** To corroborate whether vessels on the surface of the optic nerve head can provide protection against the loss of underlying axons in subjects with manifest glaucoma.

**Methods:** In this pilot study, thirty-six glaucomatous eyes with a perimetric defect in the Bjerrum area were included. The retinal nerve fiber layer (RNFL) thickness was measured in each of the sectors of the clock-hour map obtained by Cirrus HD-OCT considering the presence or absence of blood vessels. These sectors were related with their corresponding areas of the retina examined in the visual field using a mathematical model of the retina introduced by Jansonius, in order to determine the values of threshold sensitivity in those areas in the presence or absence of vessels.

**Results:** We corroborated the protective role of the blood vessel for peripapillary RNFL thickness of clock-hour 12 despite obtaining a p-value (p = 0.023; w = 228.5) close to the acceptance zone (p ≥ 0.05). The mean ± standard deviation with vessel and without vessel were 70.95 ± 24.35 and 88.46 ± 23.96, respectively. No differences were found between the mean values of threshold sensitivity to the presence or absence of blood vessels in each of the sectors considered.

**Conclusions:** Our findings do not allow us to affirm that there is an association between the presence of a vessel and protection against glaucomatous damage in subjects with an advanced manifestation of the disease. In the future, more extensive studies are needed to study this relationship in subjects with early glaucoma.

## Introduction

Glaucoma is a multifactorial progressive optic neuropathy in which elevated intraocular pressure (IOP) is a strong risk factor [1]. The structural changes that occur during the development of the disease are well-defined [2], unlike its etiology, which still remains uncertain today.

The insufficient explanation of the pathogenesis of glaucoma by mechanical and vascular theory raises interest in the permeability of the optic nerve making way for a new hypothesis that name the existence of posterior aqueous humor flow could be a possible cause of glaucoma [3]. That explains and defines the structural changes that have occurred in the development of the disease to achieve its detection as early as possible, thanks to the incorporation and improvement of imaging diagnostic devices [4-7] without having to wait to the functional manifestations of the visual field, in which case the loss of nerve fibers is irreversible and of great amount [8].

There is a fourth posterior pathway through which part of the aqueous humor leaves the eye, through the vitreous and the retina, without entering the anterior chamber. This is due to the lack of an epithelial barrier on the anterior surface of the vitreous. When there is an increase in resistance in the regulation of the aqueous humor outflow normal pathway with an increase in IOP, these discontinuities or holes produce an increase in aqueous humor flow toward the posterior pole [9]. This process is supported by the existence of optic nerve head (ONH) permeability, since the nerve’s vitreous interface is formed by astrocytes, where fenestrations are described that allow the free passage of fluid and solutes from the vitreous to prelaminar tissue [10, 11]. These astrocytes are united to each other and to the extracellular matrix, among others, by adherent junctions rich in N-cadherine, a calcium dependent adhesion molecule. These types of unions are characterized by allowing the passage of water and small molecules, which confer a permeability to the ONH [9]. When the aqueous humor, poor in calcium, passes through the optic nerve and comes into contact with astrocytes, it produces a separation of the adherent junctions due to a drop in calcium concentrations in these junctions. This would produce a rupture of the membrane junctions between the astrocytes and cause their destruction, allowing the introduction of the aqueous humor into the extracellular spaces of the optic nerve [11,12]. Do not forget that astrocytes are considered as essential elements for protecting axons of the optic nerve [3].

This new hypothesis, which is defended in the present study, should explain the loss of axons in conditions of high or low IOP and account for the initial or early damage of glaucoma in the ONH as a consequence of the deviation of the aqueous humor outlet during the development of glaucoma toward the posterior pole, in which there is protective role played by the blood vessel covering the underlying axons [3, 9].

This new postulate would allow determining the pattern of visual field loss because the scotomas would be limited in their extent by the projection of the blood vessels in the visual field [9,11,12]. Being possible to carry out a detection as early as possible, through the techniques of perimetric and morphological data, to adopt the appropriate therapeutic measures.

There are mathematical models that can be used to predict the location and progression of scotomas in glaucoma. We propose to use the Jansonius model [13,14], based on photographic images of the retina, to relate structural and functional injury. The area of the papilla covered by the central vessels of the retina corresponded to the delimited papillary border by dividing the optic nerve into sectors generated by OCT. While the Jansonius algorithm determines the area of the retina where the dendritic trees of the ganglion cells are located.

The objective of the present study is to confirm, in patients affected by glaucomatous visual field damage, whether the presence of a blood vessel on the surface of the ONH provides protection against the progressive destruction of underlying axons.

## Methods

### Study participants

This was a cross-sectional study. All subjects were recruited from the hypertension consultation at the Vista-Ircovisión ophthalmological center, in Murcia between May 2018 and September 2018.

All subjects underwent a full ophthalmic examination: a review of clinical history, best-corrected visual acuity, slit-lamp biomicroscopy, intraocular pressure measurement by applanation tonometry, automated perimetric-visual field assessment with Octopus 600 (Bloss, software version 3.6.1. Haag-Streit, Switzerland) and peripapillary analysis using optical coherence tomography (OCT) with Cirrus HD-OCT (Carl Zeiss Meditec, Inc., Dublin, CA; software version: 6.0.2.81) based on spectral domain (SD) technology using the Optic Disc Cube 200×200 protocol.

The inclusion criteria for both normal subjects and glaucoma patients were age > 18 years old, no history of retinal disease or optic nerve disease including non-glaucomatous optic neuropathy, ocular surgery, diabetes mellitus, hypertension and refractive error < 5 diopters of sphere or 3 diopters of cylinder. Glaucoma subjects were included if they had a definitive diagnosis of visual field damage in the Bjerrum area.

### Study protocol

The ONH parameters that were analyzed were RNFL thickness of all 12 clock-hour sectors listing them clockwise in the right eye and counterclockwise in the left eye. The presence or absence of a blood vessel, in each of the sectors studied in the peripapillary region, was done by superimposing the clock-hour map on the fundus photograph obtained with OCT (Fig. 1).

**Figure 1.**
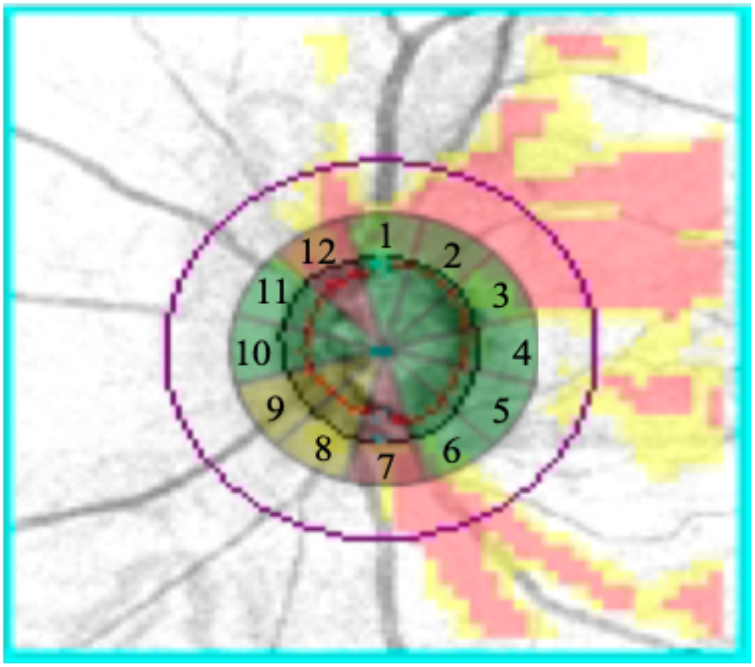
Superimposition of the clock-hour map onto the fundus photography of the optic nerve with Cirrus-HD. We observe the presence of blood vessels at clock-hour 1, 7, 8, 9 and 11 in a patient diagnosed with glaucoma.

In order to obtain threshold sensitivity values, the mathematical retinal model of Jansonius (2009) [13] was used to connect the bundle of nerve fibers of a certain region of the retina examined by the visual field with the corresponding area of input of that bundle of fibers in the peripapillary region of the optic nerve. Jansonius (2009) [13] introduced a mathematical model for describing the nerve fiber bundle trajectories of the human retina. To do this, we adapted the clock-hour map within the determined position of the optic nerve in the Jansonius model (2009) [13] matching each fiber bundle with each of the 12 sectors of the clock-hour map, thus obtaining our own proposed model to relate the structural and functional damage (Fig. 2). The threshold sensitivity value per sector was obtained by projecting each bundle of Jansonius nerve fibers onto the visual field examination considering the inversion that needed to be made (the nasal hemiretina was evaluated by the temporal visual field, the temporal hemiretina was evaluated by the nasal visual field, the superior hemiretina was evaluated by the inferior visual field and the inferior hemiretina was evaluated by the superior visual field).

**Figure 2.**
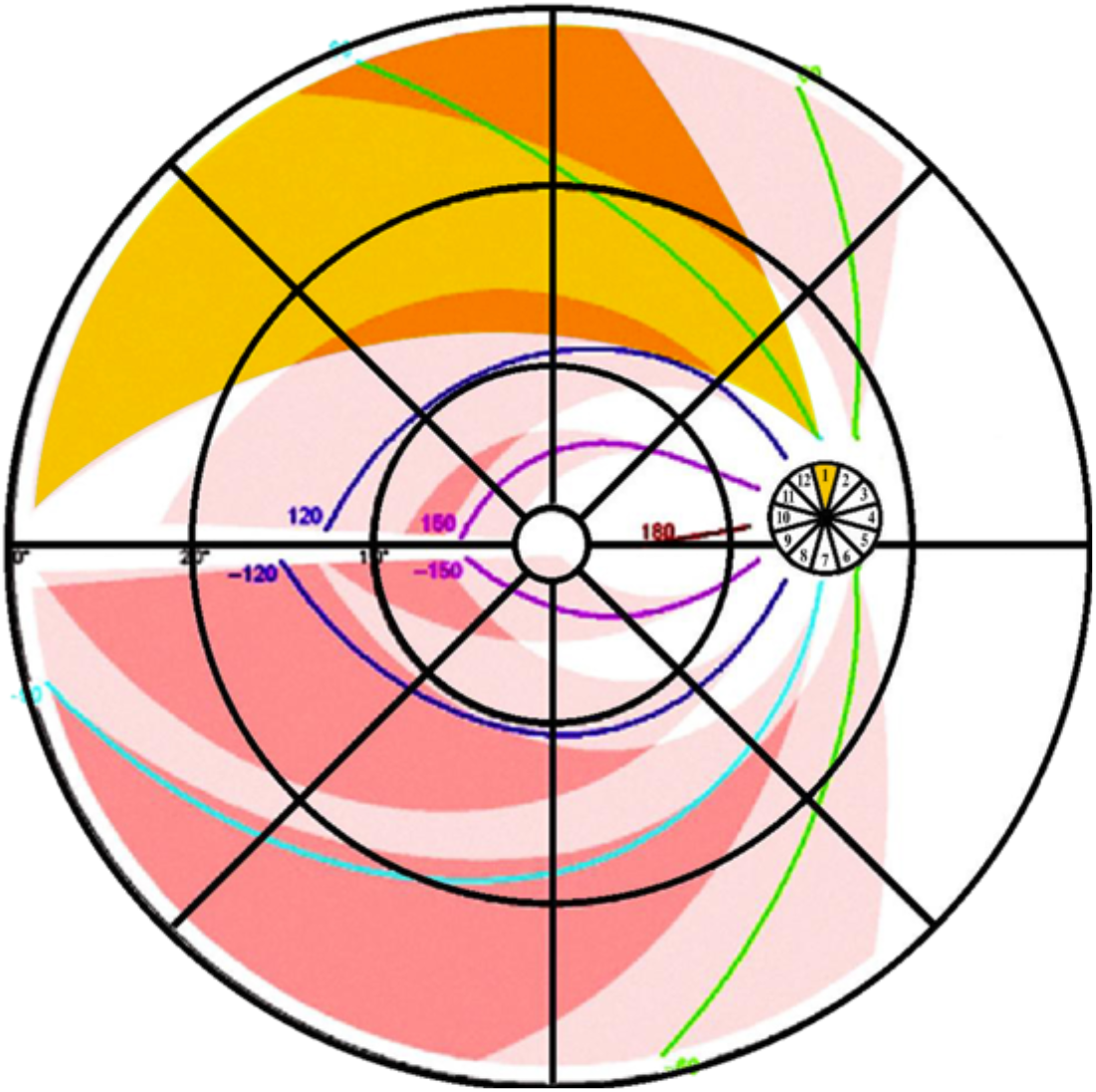
Adaptation of the clock-hour map to the mathematical retina model of Jansonius (2009). It is observed how each nerve fiber bundle trajectory in the human retina matches to the corresponding individual clock-hour sector of the ONH (the whole 12 clock-hours). An example is shown of the correspondence made in the right eye between the trajectory of the bundle axons along the retina according to Jansonius (indicated in yellow) and its corresponding input region (clock-hour 1 in yellow) in the ONH.

We calculated the mean RNFL thickness values and threshold sensitivity by stratifying each of the sectors into two groups, in order to subsequently compare the glaucomatous group with the presence of the blood vessel versus the glaucomatous group with absence of the blood vessel in the analysis of the protective role of vessels in glaucomatous damage. Similarly, previously, we measured the average values of RNFL thickness and threshold sensitivity by sectors in the recruited group of control subjects to verify the validation of our own proposed model. For this study, clock-hours 3, 4 and 5 were not included in the statistical analysis of the variables due to the limited data obtained from this area because it corresponds to the nasal side of the retina and is not affected by glaucomatous injury.

### OCT Scanning

Peripapillary ONH parameters were measured in each participant by optical coherence tomography. Optic Disc Cube 200×200 protocol of Cirrus HD-OCT was used for scan acquisition of the optical disc head. The protocol generates a cube of data over a 6×6 mm2 area centered on the optic disc by acquiring a series of 200 A-scans from 200 linear B-scans (40,000 points) in about 1.5 seconds (27,000 A-scans/sec). For analysis, a built-in automated algorithm identifies the center of the optic disc and places a circle of 3.46 mm diameter evenly around it. After automated RNFL segmentation, the system takes samples from the data cube of 256 A-scans along the path of the scan circle to obtain average and sectorial (12 clock-hours) RNFL thickness. While centering and performing the Cirrus-HD OCT test, patient monitoring is possible. Only good quality scans were used for analysis, defined as having a signal strength greater or equal to 6 (10 = maximum) without discontinuity or misalignment, involuntary saccade or blinking artifacts. The parameters used for the analysis of the optic nerve head were the clock-hour map and mean nerve fiber layer (NFL) thickness.

### Visual Field Testing

All participants underwent the G-Program of the Octopus perimeter, TOP strategy, 30 central degrees, white-on-white perimetry, stimulus size III, time of exposure 100ms using standard background lighting of 4 apostilbs. The visual fields were classified as reliable if the reliability factor, RF, was <15%.

Glaucomatous visual field damage was defined based on at least 3 or more contiguous depressive test points within the typical glaucoma location (Bjerrum area) with a pattern deviation plot at p<5% or a corrected standard deviation occurring in less than 5% of the normal field.

### Statistical analysis

To compare the means in the different groups, the Wilcoxon-Mann-Whitney test was used. R Studio version 1.1.423 was used for statistical analysis in order to have a simple way to execute the sentences of the programming language R version 3.5.0 (2018-04-23) necessary to obtain the desired analyses. In all analyses, p-values < 0.05 were considered significant.

## Results

The following participants were recruited: 27 control subjects (36 eyes) and 29 patients (36 eyes) diagnosed with primary open-angle glaucoma (POAG). In all, 72 patients aged 50-79 years were included. The patients of the study group had been diagnosed by a glaucoma specialist and were in follow-up for the disease, after having undergone the functional test with at least 5 visual fields prior to their recruitment. Even so, 9 patients were eliminated because the reliability limits established in the perimeter used were exceeded (RF <15%). All patients had glaucomatous visual field defects (Bjerrum area) and glaucoma was classified as advanced if the average value of MD = 10.25 ± 4.98 (>/= 6dB). The normal control subjects presented normal perimetries and no evidence of structural or functional glaucomatous damage (Table 1).

**Table 1.**
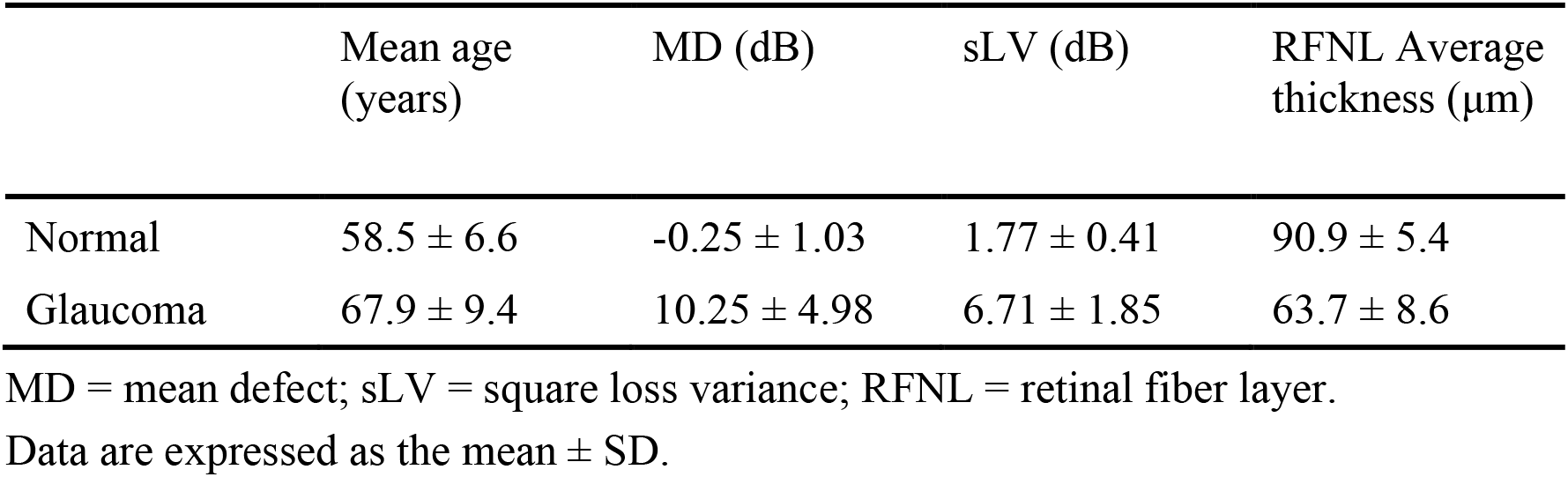
Baseline characteristics of the normal control and glaucoma patients.

Regarding our work methodology, we found our regions of the visual field with their corresponding sections of analysis of nerve fibers of the ONH of the hour circle by OCT, when comparing our model with that used in the only study found among the bibliography that performs a topographical correspondence between functional and structural tests [15]. No other Jansonius-based model was found. Likewise, we found analogous with other studies the mean values of nerve fiber thickness in each of the sections considered (Table 2), [5,16] as well as the value of the mean visual field defect [17] both in the group of subjects. control as glaucomatous patients.

**Table 2.** Peripapillary Retinal Nerve Fiber Layer Thickness (μm) of normal control and glaucoma patients by location for Cirrus HD-OCT.

| Parameter | Normal | Glaucoma |
| --- | --- | --- |
| Sector 1 | $111.91 \pm 22.22$ | $72.72 \pm 19.90$ |
| Sector 2 | $102.33 \pm 13.00$ | $74.11 \pm 18.82$ |
| Sector 6 | $93.77 \pm 16.09$ | $66.88 \pm 14.93$ |
| Sector 7 | $125.69 \pm 17.34$ | $73.80 \pm 18.94$ |
| Sector 8 | $131.58 \pm 16.84$ | $68.41 \pm 19.16$ |
| Sector 9 | $63.38 \pm 9.45$ | $47.27 \pm 11.64$ |
| Sector 10 | $49.16 \pm 5.16$ | $47.36 \pm 12.07$ |
| Sector 11 | $73.86 \pm 9.97$ | $58.00 \pm 16.07$ |
| Sector 12 | $126.05 \pm 15.05$ | $78.25 \pm 25.40$ |

Data are expressed as the mean ± SD

We can affirm that our data obtained from our analyzed sample are correct when finding common analogous findings with respect to other studies. This, together with the previous knowledge we have of the disease, allows us to accept as valid our proposed model to match the regions of the visual field with their corresponding sectors of the optic nerve.

Therefore, we start from the creation of a correct model that allows us to study / analyze the main objective of this work, which is to corroborate if the blood vessel really has a protective role against glaucomatous lesion. We found that its influence was only confirmed in section 12 of the examination of the hourly circle for the thickness of NFL after the application of the WMW test, after checking the non-normality of the variables involved using the Shapiro-Wilk test, confirming the rejection of equality of means (μ1 = μ2) despite the fact that the obtained value was very close to the acceptance zone (p ≥ 0.05), considering therefore the test that for this section the vessel does influence the thickness of NFL in glaucomatous subjects (Table 3).

**Table 3.**
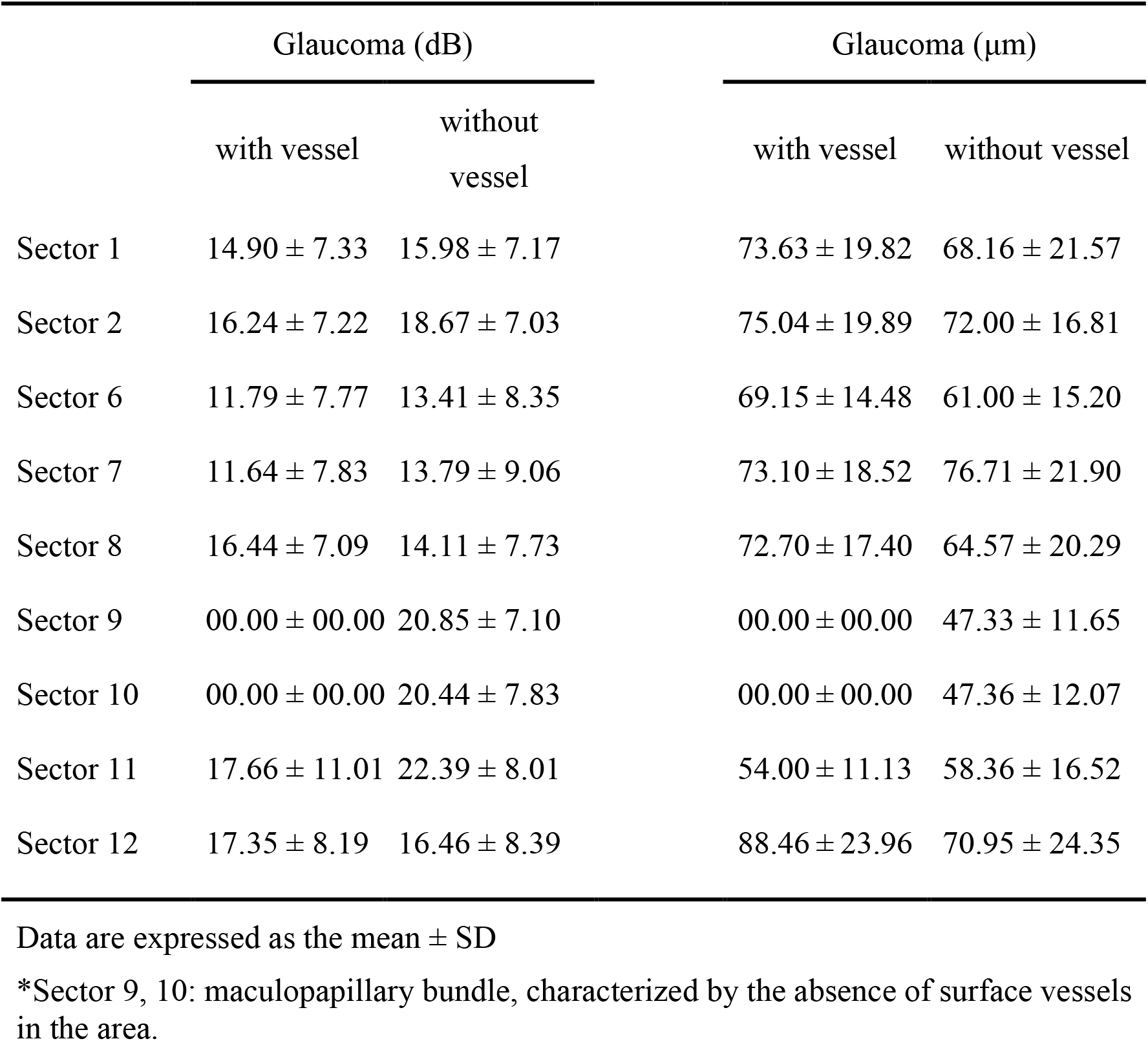
Comparison of results of mean sensitivity (dB) and peripapillary RNFL (μm) of glaucoma patients by location with vessel/without vessel.

## Discussion

The application of an early diagnosis of glaucoma and early intervention treatment plan, without having to wait for clearly defined visual field losses, has led to the emergence of a variety of types of new diagnostic imaging technology with the objective of measuring structural changes which is very useful in glaucoma treatment [4-7].

In spite of all the studies that have attempted to confirm that the new imaging techniques are good predictors of early structural glaucomatous damage, others question the ability of these techniques [18–21].

There is no unanimous consensus on whether structural damage precedes functional progression in glaucomatous eyes [22, 23]. It is probably caused by a definition problem due to the limitation of instrumental capacity, specificity and sensitivity [24]. In addition, we must not forget that we define and understand the term “early diagnosis” as the presence of early signs of glaucoma, which in turn are characterized by being highly variable and difficult to catalog [25]. In the clinical setting, we often find patients with evidence of glaucomatous optic neuropathy without a detectable visual field abnormality, patients with a glaucomatous visual field without detectable structural abnormality and patients with a similar degree of stages of glaucomatous structural alterations but with different levels of losses in the field visual [22]. Therefore, the diagnosis and monitoring of glaucoma cannot be made based on a single examination.

We believe, like other researchers, that the problem really lies in the difficulty of the concordance and relationship between the perception of the differential threshold obtained by the visual field and the reduction of ganglion cells detected by the OCT [26]. The problem with all this lies in the fact that there is no linear relationship between the measurements used in each test. The visual field uses logarithmic units of measurement, decibels (dB), measuring the differential light sensitivity of the subject which are related to the luminance of the stimulus relative to the background. Therefore, it does not express the value of the detected luminosity as an absolute value but as a relative value. In addition, OCT measures the thickness of peripapillary RNFL in linear units (μm) giving a reference value with respect to normality measurements, as the μm is a linear unit which has a direct measurement relationship with retinal ganglion cell density [22, 24].

It has been tried, without success, to create a model that relates both tests. Several authors have attempted to do this, and even here there is still a diversity of opinions. Some believe it is better to generate models whose association is considered more correct if both tests are transferred to linear units [27] and others believe that, on the contrary, the association is better if it is done in logarithmic units [29].

A possible solution and contribution, particularly for the improvement of lineal models, is the model we propose in our study. We suggest determining a map in order to make up for the lack of knowledge, which determines a relationship between structure and functional damage in glaucoma, to match a bundle of nerve fibers of a certain region of the retina examined by the visual field with the corresponding area of input of that bundle of fibers in its corresponding sector in the ONH. In this way, we could obtain the thickness of peripapillary nerve fibers measured by the OCT and the sensitivity values measured by the visual field corresponding to each clock-hour sector. The best-considered and most-used model for this is the Galway Heath (2000) [28] on which the Jansonius retinotopic model (2009) [13] is based, and which we have used to generate our own model. It differs from the rest by making a mathematical model mapping nerve fiber bundle trayectories in the retina, describing a clear asymmetry between the superior and inferior hemifields and providing a detailed location-specific estimate of the magnitude of variability [14].

According to our data (Table 2), we can affirm that our model, which relates structural to functional function, is reliable in trying to respond to the objective of this study since all efforts are directed towards early detection of the disease in order to thus approaching it with the most appropriate therapeutic considerations [25]. Reflection that leads us to ask ourselves how to know that therapeutic action to apply is the correct one if the etiopathogenesis of glaucoma is not really known. To resolve this question, after initially programming our work methodology, we decided to clinically check one of the postulates that this new theory defends [30,31]: if the blood vessel really has a protective role against glaucomatous lesion both at the level of fiber thickness nerve as in light sensitivity values in each of the 9 sections considered in the groups formed (group of subjects with glaucoma in whose sections there is a presence of a vessel versus group of subjects with glaucoma in whose sections there is no presence of a vessel) (Table 3). Note that the sections in which we are going to find the position of a vessel will be section 1 and 7, followed by section 2 and 6, respectively corresponding to the limiting region of the fibers of the Bjerrum area and immediately more peripheral to these. In contrast, the sections belonging to the papillo-macular bundle (sections 9, 10 and 11) are the most numerous in the absence of a blood vessel.

## Conclusion

In conclusion, we only found the protective role of a blood vessel was strongest for clock-hour 12 on the thickness RFNL. We did not find an influence of the protective role of a blood vessel for sensitivity values in any individual sector of the clock-hour map studied. We cannot confirm that the blood vessel has a protective role against glaucomatous damage based on the data obtained and we are not able to compare our results with those obtained by other authors. To our knowledge, this is the first study that clinically proves the protective role hypothesis of the blood vessel in glaucomatous damage and it supports new theories that may determine the pathogenesis of glaucoma.

Based on these findings, additional studies are needed to evaluate the protective role of the blood vessel applied to subjects with early glaucomatous damage, to check whether there is a greater advance in the disease comparing those sectors with or without vascular protection. It would also be interesting to study functionality by microperimetry which has the capacity to detect minimum variations of the visual threshold in a more precise way than the visual field.

## Data Availability

The datasets used and/or analyzed during the current study are available from the corresponding author on reasonable request.

## Abbreviations

IOP: Intraocular pressure
ONH: Optic nerve head
POAG: Primary open angle glaucoma
OCT: Optical coherence tomography
SD: Spectral domain
RNFL: Retinal nerve fiber layer
NFL: Nerve fiber layer.

## Declarations

### Conflicts of interests

The authors declares that there is no conflict of interest regarding the publication of this article.

### Funding Statement

No funding was received for this research.

## Acknowledgements

Not applicable.

## Authors’ contributions

YC: data acquisition and analysis, interpretation of data, drafting the manuscript and critical revision of the manuscript. JL: research design and critical revision of the manuscript. LV, MR, JL: critical revision of the manuscript. RL: analysis and interpretation of data. All authors have read and approved the manuscript.

## Approved by the following research ethics committee

This study was approved by the Ethics Committee of the Catholic University of Murcia (CE041809). The present investigation was conducted according to the tenets of the Declaration of Helsinki and current regulations to protect the confidentiality of data. The research is based entirely on the clinical routine and is obtained from the patient’s medical record system. Therefore, no informed consent is required.

## Consent for publication

Not applicable

## Additional file

STROBE Statement—Checklist of items that should be included in reports of ***cross-sectional studies***

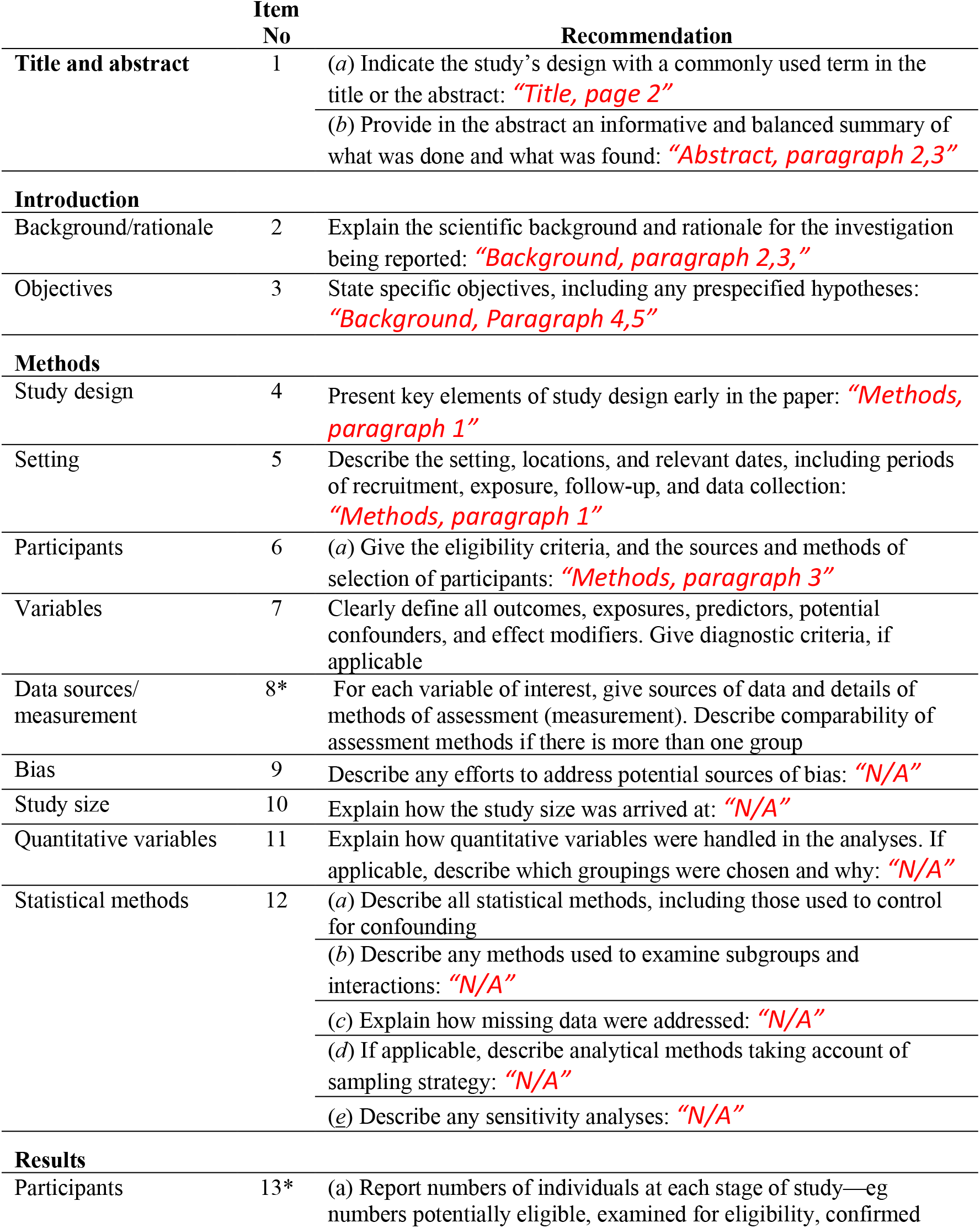
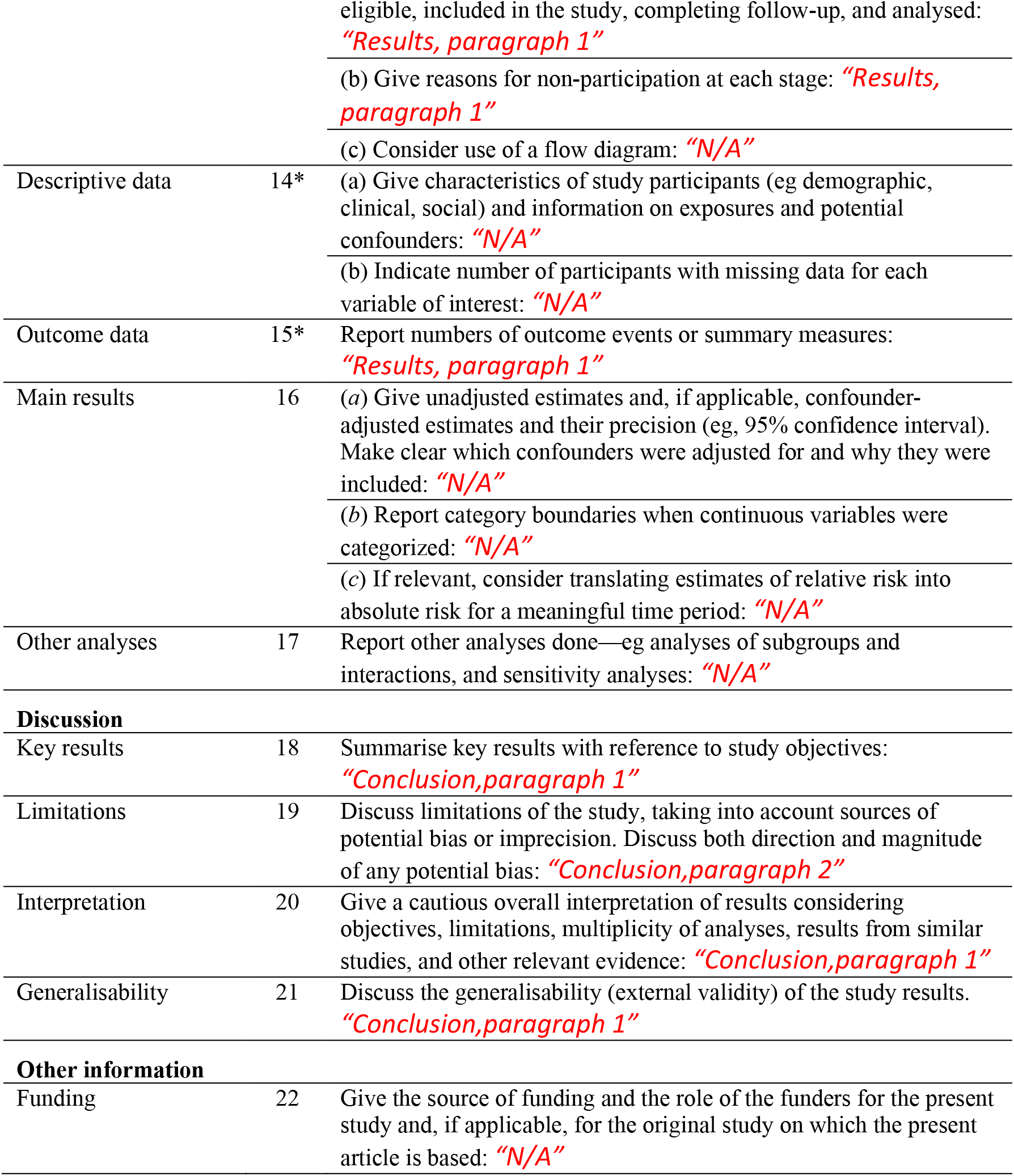

## Notes

### Competing Interest Statement

The authors have declared no competing interest.

### Author Declarations

This study was approved by the Ethics Committee of the Catholic University of Murcia (CE041809). The present investigation was conducted according to the tenets of the Declaration of Helsinki and current regulations to protect the confidentiality of data. The research is based entirely on the clinical routine and is obtained from the patients medical record system. Therefore, no informed consent is required.

